# The Best Start (Kia Tīmata Pai): A Study Protocol for a Cluster Randomized Trial with Early Childhood Teachers to Support Children’s Oral Language and Self-Regulation Development

**DOI:** 10.1101/2022.10.18.22281232

**Authors:** Elaine Reese, Jesse Kokaua, Hayley Guiney, Tugce Bakir-Demir, Jimmy McLauchlan, Clair Edgeler, Elizabeth Schaughency, Mele Taumoepeau, Karen Salmon, Amanda Clifford, Natasha Maruariki, Stuart McNaughton, Peter Gluckman, Charles A. Nelson, Justin M. O’Sullivan, Ran Wei, Valentina Pergher, Sophia Amjad, Anita Trudgen, Richie Poulton

## Abstract

**Introduction:** Oral language skills are associated with children’s later self-regulation and academic skills; in turn, self-regulation in early childhood predicts successful functioning later in life. The primary objective of this study is to evaluate the separate and combined effectiveness of an oral language intervention (ENRICH) and a self-regulation intervention (ENGAGE) with early childhood teachers and parents for children’s oral language, self-regulation, and academic functioning.

**Methods and Analysis:** The Best Start study (Kia Tīmata Pai in te reo Māori, the indigenous language of New Zealand) is a cluster randomized controlled trial with teachers and children in approximately 140 early childhood centres in New Zealand. Centres are randomly assigned to receive either oral language intervention only (ENRICH), self-regulation intervention only (ENGAGE), both interventions (ENRICH + ENGAGE), or an active control condition. Teachers’ and parents’ practices and children’s oral language and self-regulation development are assessed at baseline at age 1.5 years and every 9-months to age 5, and academic performance at age 6. Teacher-child interactions will also be videotaped each year in a subset of the centres. Children’s brain and behavior development and parent-child interactions will be assessed every 6 months to age 6 years in a sub-group of volunteers.

**Ethics and Dissemination:** The Best Start trial and the two sub-studies (Video Project; Brain and Behavior Development) have been approved by the University of Otago Health Ethics Committee (H20/116), and reviewed for cultural responsiveness by: the Ngāi Tahu Research Committee (University of Otago), the Māori Advisory Group (University of Auckland, Liggins Institute) and an internal cultural advisory group. Results will be disseminated in international and national peer-reviewed academic journals and communicated to local, national, and international organizations serving early childhood teachers, parents, and young children.

**Trial registration:** This trial is registered with the Australian New Zealand Clinical Trial Registry (ANZCTR) as ACTRN12621000845831.

**Strengths and Limitations of This Study:**

- **Strength:** The size and longitudinal design of this cluster RCT across early childhood will enable the assessment of the singular and combined effects of teacher-led oral language and self-regulation interventions on the development of children’s oral language, self-regulation, and academic functioning.
- **Strength:** The trial represents a partnership between a large provider of early childhood education (BestStart), an external implementation service (Methodist Mission Southern), and a consortium of academics (Emotion Regulation Aoetaroa/New Zealand) to provide a culturally responsive intervention with co-designed implementation with the early childhood education provider (BestStart); the use of a single national provider simplifies co-design and the implementation of the intervention.
- **Strength:** The efficacy of the interventions is assessed at multiple levels: via teacher and parent reports, behavioral observations, and measures of brain development (EEG/ERP).
- **Limitation:** Because of the large size of the main trial, those assessments are restricted to teacher- and parent-report instruments.
- **Limitation:** The single-provider feature with BestStart restricts the ability to collect primary outcomes on children who leave this service provider during early childhood.

## Introduction

Self-regulation comprises the ability to regulate one’s thoughts, feelings, and actions [1]. Self-regulation skills are developing rapidly in early childhood [2] and predict later academic functioning and life success [1, 3]. Clinically relevant difficulties in these skills predict negative outcomes across domains of functioning, including psychological and physical health across the lifespan [4]. Self-regulation difficulties are linked to later problems in academic, occupational, and socioemotional functioning [5-7]. Therefore, it is vital to design preventive measures that can be implemented during early childhood when these skills are rapidly developing.

A successful play-based preventive intervention called ENGAGE (Enhancing Neurocognitive Growth with the Aid of Games and Exercise) [8, 9] aims to improve children’s cognitive, emotional, and behavioral self-regulation through children’s games (e.g., Simon Says) that are interpersonal in nature and teach a range of skills. ENGAGE leads to improvements in parent-rated behavior problems equivalent to a gold-standard parent-management programme (Triple P), with treatment gains maintained 12 months later [9]. In an intervention in 28 early childhood education centres (ECEs) in Auckland New Zealand, ENGAGE was demonstrated to lower levels of hyperactivity, aggression, and inattention across a cohort of 940 preschool children relative to a wait-list control period [10].

Self-regulation can be also be fostered by enhancing children’s oral language development [11]. The way adults talk with children during everyday activities (book-reading, mealtimes, play) advances children’s early language and cognitive development [12], which in turn enhances their self-regulation [13]. Book-sharing [14] and conversing about everyday experiences [15] are two evidence-based methods that improve children’s oral language skills. For instance, a combined reading-and-conversation program developed in New Zealand with parents and teachers called *Tender Shoots* improves preschool children’s oral narrative, early literacy, and socioemotional skills up to one year after the intervention when compared to activity-based controls [16-19]. Critically, children’s academic skills at school entry predict their later academic and socioemotional functioning [20].

Oral language is developing rapidly in the toddler years. Therefore, oral language interventions at this age can produce stronger benefits than similar interventions at older ages [21], but such interventions are rare [13]. Previous oral language interventions have focused primarily on training either parents or teachers to engage in the interactions, but not both [22]. The few combined interventions that have been conducted indicate larger benefits for preschool children when both parents and teachers are using the techniques compared to home-only or school-only [23, 24]. We have developed a shared reading-and-conversation program for even younger children with both teachers and parents called ENRICH (Enhancing Rich Conversations; see Table 1 and Figure 1).

**Table 1.** Books and Cards for ENRICH and ENRICH+ Interventions

|  | *ENRICH (1.5 to 3 years)* | *ENRICH+ (3 to 5 years)* |
| --- | --- | --- |
| *Phase 1 Books* | ➤ *The Noisy Book* by Soledad Bravi<br>➤ *Hoihoi Turituri* transl. by Ruia Aperahama | ➤ *Pukeko Shoes* by Janet Martin and Ivar Treskon<br>➤ *The Little Red Hen* by Paul Galdone<br>➤ *Cuddly Dudley* by Jez Alborough<br>➤ *Five Little Monkeys Sitting in a Tree* by Eileen Christelow |
| *Phase 1 Cards* | ➤ Overview of ENRICH, Phase 1<br>➤ Kai Time<br>➤ Book Time<br>➤ Nappy Time<br>➤ Play Time<br>➤ Group Time | ➤ Overview of ENRICH+, Phase 1<br>➤ Kai Time<br>➤ Book Time<br>➤ Greetings/Farewells<br>➤ Play Time<br>➤ Group Time |
| *Phase 2 Books* | ➤ *Mahi/Actions* by Kitty Brown and Kirsten Parkinson<br>➤ *Kare ā-roto/Feelings* by Kitty Brown and Kirsten Parkinson<br>➤ *Who's Driving?* by Leo Timmers<br>➤ *Ma Wai e Hautu* transl. by Karena Kelly | ➤ *The Gingerbread Man* by Eric A. Kimmel<br>➤ *Bunny Cakes* by Rosemary Wells<br>➤ *Louie the Tui* by Janet Martin<br>➤ *Elephant Island* by Leo Timmers |
| *Phase 2 Cards* | ➤ Overview of ENRICH, Phase 2<br>➤ Kai Time<br>➤ Book Time<br>➤ Nappy/Toilet Time<br>➤ Play Time<br>➤ Group Time | ➤ Overview of ENRICH+, Phase 2<br>➤ Kai Time<br>➤ Book Time<br>➤ Greetings/Farewells<br>➤ Play Time<br>➤ Group Time |
| *Phase 3 Books* |  | ➤ *3 Billy Goats Gruff* by Paul Galdone<br>➤ *Slinky Malinki Early Bird* by Lynley Dodd<br>➤ *That's Not a Hippopotamus!* by Juliette MacIver and Sarah Davis<br>➤ *Jumblebum* by Chae Strathie and Ben Cort |
| *Phase 3 Cards* |  | ➤ Overview of ENRICH+, Phase 3<br>➤ Kai Time<br>➤ Book Time<br>➤ Greetings/Farewells<br>➤ Play Time<br>➤ Group Time |

**Figure 1.**
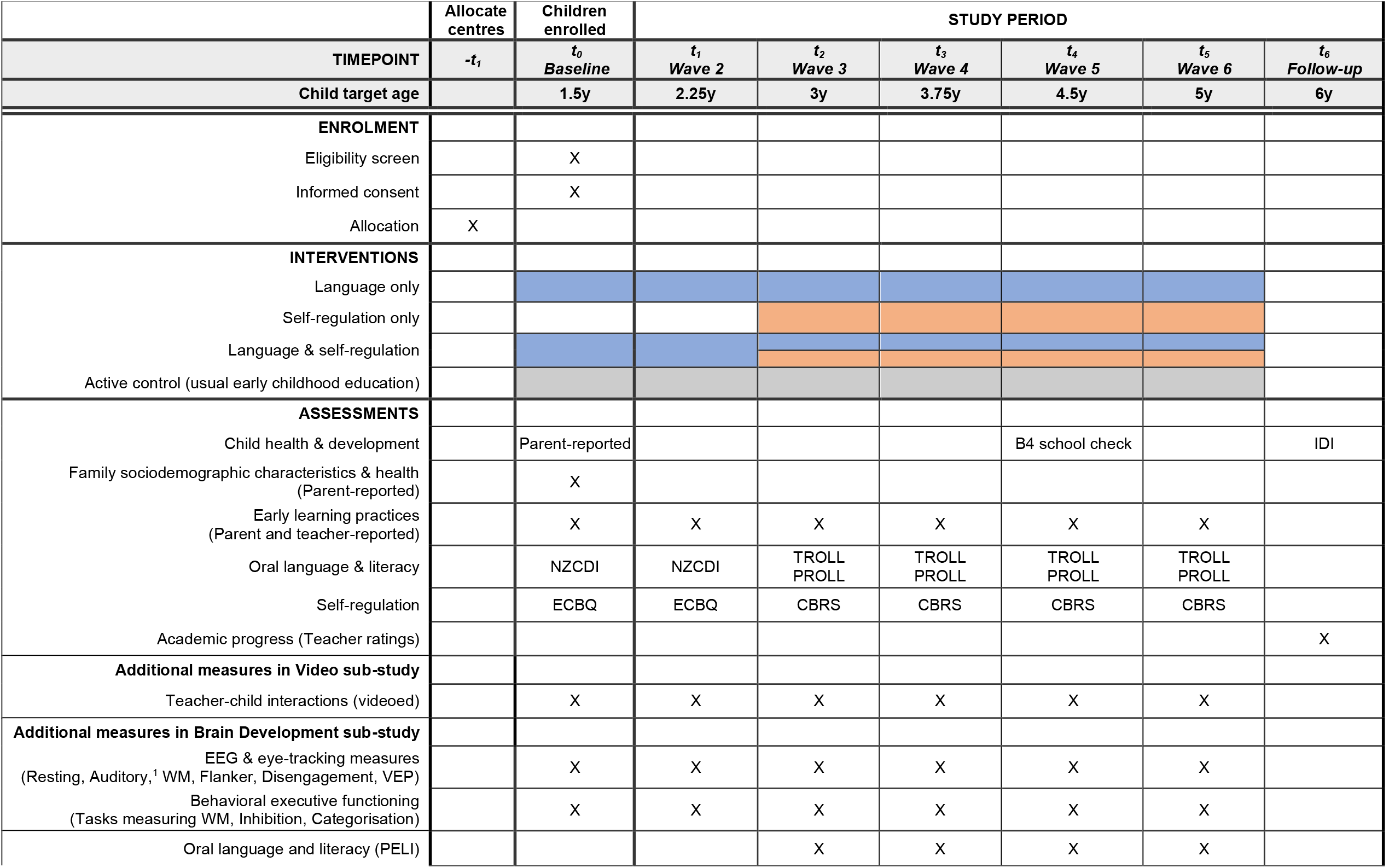
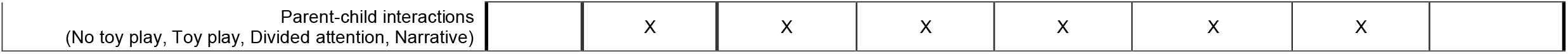
Kia Tīmata Pai schedule of enrolment, interventions, and assessments. *Notes*. Specific measures are listed at a particular time-point if there is change in the measure used across time due to ensure they remain developmentally-appropriate. ‘X’ indicates that a particular set of measures was used at that time point. ^1^EEG only. ECE = early childhood education. NZCDI = New Zealand Communicative Development Inventories. TROLL = Teacher Rating of Oral Language and Literacy. PROLL = Parent Rating of Oral Language and Literacy. ECBQ = Early Childhood Behaviour Questionnaire (surgency, negative affect, and effortful control subscales). CBRS = Child Behavior Rating Scale. IDI = Integrated Data Infrastructure. WM = working memory. VEP = visually-evoked potential. PELI = Preschool Early Literacy Indicators. Narrative = shared past event narrative.

### Objective

To determine if an intervention, over and above the usual early childhood curriculum, that targets development of oral language plus self-regulation (ENRICH plus ENGAGE) has greater benefits on the development of children’s oral language, self-regulation, and academic functioning than interventions that target either oral language (ENRICH), or self-regulation (ENGAGE).

## Methods and Analysis

### Study design and participants

This study is an open-label, cluster designed four-armed randomized controlled trial (RCT) with children aged 13 to 30 months at outset, their parents, and their teachers at BestStart early childhood centres in New Zealand. ECEs are randomly assigned to the ENRICH (Enhancing RICH Interactions) and/or ENGAGE (Enhancing Neurocognitive Growth with the Aid of Games and Exercise) interventions or an Active Control group; teacher and family participation in the study is voluntary at all stages. Interventions are implemented with a factorial design.

Professional development with teachers and workshops with parents began when children were 13 to 30 months (baseline). Interventions will continue until children are 5 years of age, and children will be followed until they are 6 years of age (see Figures 1 and 2; supplemental file 1). The ENRICH intervention is book- and conversation-based to foster toddlers’ oral language and literacy skills, with ENRICH+ an advanced version for preschoolers (see Table 1 and supplemental file 1). The ENGAGE intervention is games-based to foster preschool children’s self-regulation skills (see Table 2 and supplemental file 1). Recall that oral language is a pathway to self-regulation [11].

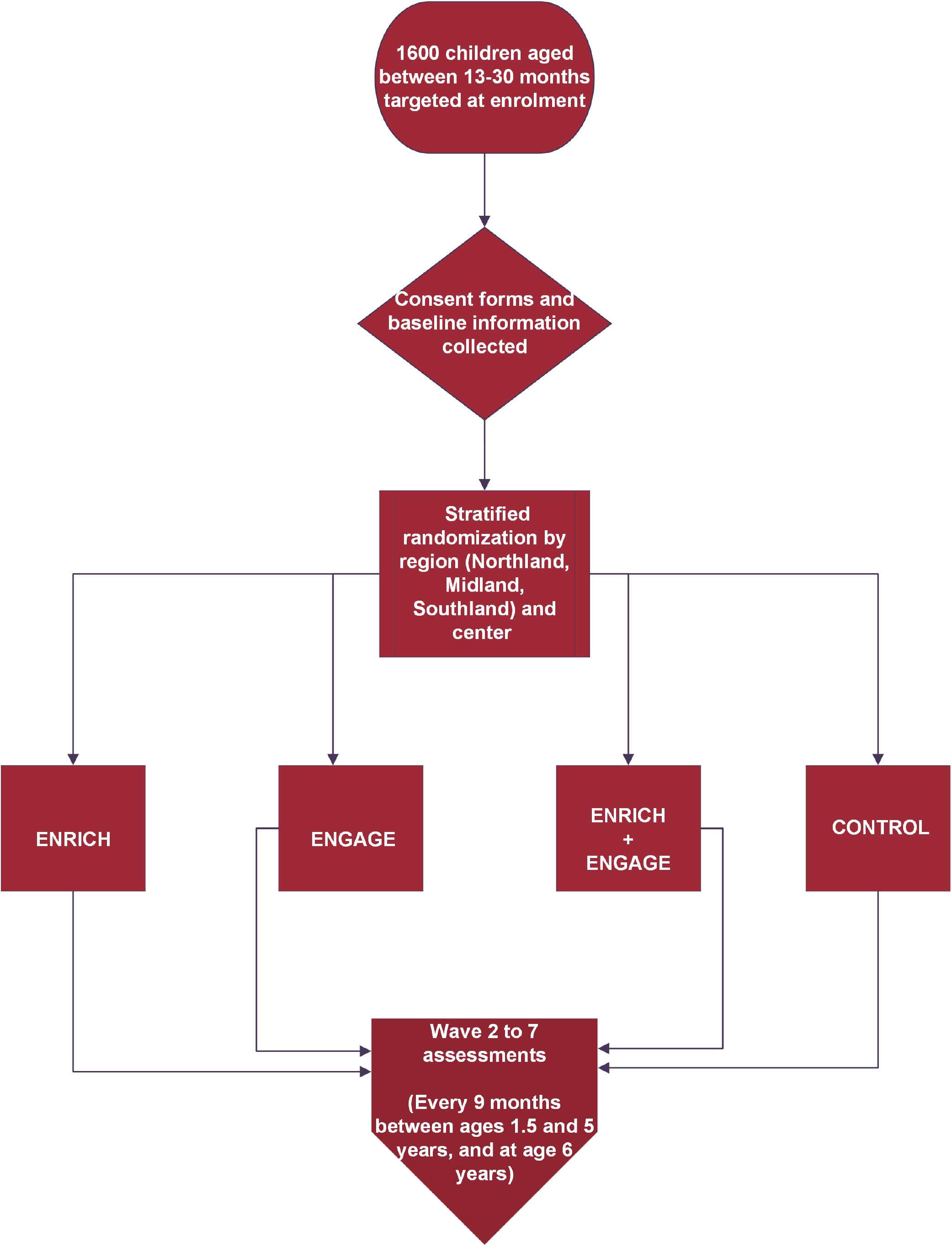

**Table 2*.** Examples of ENGAGE Games and Activities

| *Targeted Domain* | *ENGAGE Game or Activity (3 to 5 years)* |
| --- | --- |
| *Emotional/Feelings* | ➤ Relaxing Yoga<br>➤ Deep Breathing<br>➤ Drawing |
| *Cognitive/Thinking* | ➤ Object Copy<br>➤ Puzzles<br>➤ Cups Memory<br>➤ Beading<br>➤ Snap<br>➤ Card Memory |
| *Behavioral/Doing* | ➤ Ball Games<br>➤ Musical Statues<br>➤ Animal Speeds<br>➤ Skipping<br>➤ Ball and Spoon Race<br>➤ Simon Says<br>➤ Hop Scotch |

\*Adapted with permission from Healey and Healey (2019; Table 1)

ECEs, and participants, will be randomized into the following four arms:

1. Language only: ENRICH (1.5 to 3 years) and ENRICH+ (3 to 5 years)
2. Self-regulation only: ENGAGE (3 to 5 years)
3. Combined: ENRICH (1.5 to 3 years) and ENRICH+ (3 to 5 years) and ENGAGE (3 to 5 years)
4. Active Control: Curriculum as usual plus child development webinars

The Best Start trial contains two nested sub-studies: 1) the Video Project: a video teacher-child interaction sub-study to measure implementation within 24 ECE centres randomly selected from participating centres in two large cities (Auckland: 12; Christchurch; 8), two small cities (Dunedin: 1; Invercargill: 2), and a rural region (1); and 2) the Brain and Behavior Development sub-study: a study on a subset of children (n= 237) who volunteered to participate in additional neurophysiological and behavioral testing at university laboratories in either Auckland or Christchurch, New Zealand.

Teachers are being trained in the interventions using a “train the trainer” model. Firstly, teacher managers (called professional practice leaders) within the BestStart early childhood organisation are being trained online and in person in the implementation of the ENRICH and ENGAGE interventions by the research team. These professional practice leaders have at least 10 years of experience in early childhood education. The professional practice leaders subsequently conduct training workshops with the ECE teachers, again via online and in-personal delivery, who then implement the techniques with children in their centres.

Professional development workshops for the toddler phase of ENRICH (from approximately 1.5 to 3 years of age) were delivered in 2021 and 2022. Professional development workshops for ENRICH+ and ENGAGE will take place in early 2023 when study children are approximately 3 years old (i.e. at the start of the preschool phase, which extends from approximately 3 – 5 years of age).

Professional practice leaders will receive booster training sessions approximately every 9 months throughout the course of the study; these leaders will then deliver booster training sessions to teachers. As part of the study, professional practice leaders are required to document the delivery of teacher workshops and follow-ups with centres to answer questions. Every time teachers in either ENRICH or ENGAGE conditions receive new training, parents of children in intervention conditions are invited to learn about the new techniques through online information evenings and recorded video links.

Teachers in ENRICH centres were trained in 2021 and 2022 and provided with new resources for the toddler phase of the study (two sets of informational cards, 6 new books, and instructional videos; see Table 1). In the preschool phase of the study, teachers in both ENRICH+ and ENGAGE conditions will receive new training and resources (sets of informational cards and/or books and instructional videos) approximately every 9 months (see Table 1).

Teachers in ENRICH centres are encouraged to use the intervention techniques as often as possible throughout the day during five routines: mealtimes, book times, diaper changes, play, and group time. Provided books are all commercially available, but contain conversation prompts on each page that have been specially designed and inserted for this study [18]. In ENRICH+, teachers will be encouraged to select two books per week to read three times each, plus to continue to have enriched conversations on a daily basis during greetings/farewells, mealtime, play time, and group time. Teachers in ENGAGE centres are encouraged to use the games for 30 minutes a day.

In the toddler phase of the study (1.5 to 3 years), teachers in centers in the ENGAGE-only arm and the Active Control arm participated in two webinars on childhood nutrition, a topic chosen in consultation with BestStart. In the preschool phase, teachers in the Active Control arm will also receive child development webinars every 9 months on children’s friendships, a topic chosen in consultation with BestStart. All webinars are recorded and delivered by Ph.D.-level child development experts for approximately 1 hour each including questions. Parents of children in the ENGAGE-only arm (toddler phase) and the active-control arm of the study are invited to these webinars or to watch recordings, at their leisure.

### Participants: Eligibility criteria, study setting, and consent process

The Best Start | Kia Tīmata Pai trial is a collaboration with a large national early childhood education organisation in New Zealand called BestStart. In 2021 and 2022, managers from 140 ECE centres invited teachers and parents of all children in the target age range (originally from 17-24 months but changed to 13-30 months at enrolment) to consent to participate via an online link or paper forms at the centre (see supplemental file 3). There are no gender, ethnic, language, or socioeconomic restrictions to participation. All enrolled children (13 to 30 months at enrolment; *M* = 20.6 months; *SD* = 3.4) are being exposed to the interventions within their early childhood education centre. Data are being collected only on those children whose families granted consent.

### Implementation checks

Intervention fidelity is being monitored: 1) via fortnightly teacher self-ratings of the frequency of their delivery of the techniques, and each study child’s engagement in the techniques; and 2) via videotaped teacher-child interactions once a year in the 24 centres in the Video Project (approximately 100 children and their teachers each year). These implementation checks will serve as an audit on trial conduct and are being overseen by Methodist Mission Southern, a co-investigator body that is independent of the sponsoring organization (ERANZ).

### Outcomes

#### Primary

Our primary outcomes are children’s oral language, self-regulation, and early literacy (see Figure 1). In the toddler phase (1.5 years to 2.25 years), we are measuring increases in vocabulary size and syntax with the New Zealand Communicative Development Inventories (short forms) for parents and teachers [25] and increases in the effortful control subscale and decreases in the negative emotionality subscale of the Early Childhood Behavior Questionnaire [26]. In the preschool phase (3 to 5 years), we will measure increases in oral language (semantic, phonological, syntactic, and pragmatic) and literacy (letter recognition, print concepts, and writing) with the Teacher Rating of Oral Language and Literacy [27] and the adapted companion Parent Rating of Oral Language and Literacy [28]. We will measure increases in self-regulation outcomes from ages 3 to 5 with teacher and parent versions of the Child Behavior Rating Scale [29]. Finally, global teacher ratings will assess improvement of children’s oral language, literacy, self-regulation, and social skills from ages 3 to 6 [19].

#### Secondary

Children’s B4 school check at age 4.5 years will be accessed through the Ministry of Health to assess improvements in school readiness [30]. Medical practitioners rate children’s cognitive, physical, and socioemotional development. Children’s Integrated Data Infrastructure (IDI) will also be accessed at age 6 years and beyond to provide long-term data on improvements in education, health, and employment outcomes. The IDI is administrative data held by the New Zealand government on a range of outcomes.

Parents and teachers are self-reporting on their early learning practices at each assessment wave. Teachers are also reporting on their proficiency in te reo Māori, an official language of New Zealand and a target focus of some of the ENRICH materials (see Table 1).

Each year, approximately 100 children and their teachers in the Video Project classrooms are being videotaped for a total of 25 minutes (5 minutes per targeted routine). The videotapes will be transcribed and then coded for the quantity and quality of speech by coders who are blind to centre condition. Two waves of video observation data were collected in 2021 and 2022, with two more waves planned in 2023 and 2024.

Every 6 months, the children participating in the Brain and Behavior Development sub-study are being administered additional neurophysiological measures (EEG/ERP and eye-tracking), behavioral measures of executive functioning (working memory, inhibition, and categorization), oral language and literacy (vocabulary, story comprehension, phonological awareness, and letter recognition); and a parent-child interaction measure (see Figure 1 and supplemental file 2). The first wave of data is now complete, with the second and third waves ongoing, and a fourth wave planned to begin in 2023.

### Participant and public involvement

A key feature of the intervention is to co-design implementation with BestStart professional practice leaders. Academics designed the two interventions, but the way the techniques are presented to teachers at each phase is being developed through a co-design process with BestStart leaders, facilitated by implementation leaders at Methodist Mission Southern. The objective is to build on teachers’ existing knowledge and practices to enhance uptake of ENRICH and ENGAGE techniques. BestStart leaders also inform the administration of the fortnightly implementation checks and assessments at each wave.

### Sample size calculation

Based on previous oral language and self-regulation interventions with early childhood teachers [10, 13, 24, 31], we anticipate small to medium effect sizes. Power analyses of final-phase primary outcomes suggested a total sample size at baseline of approximately 1600 children (i.e., approximately 400 per group, including control group) for the current study to afford precision for estimates with alpha = 0.05 and power between .75 and .90. This sample size of 400 per group at baseline allows for attrition of children over the course of the study. This sample size also takes into account multiple predictor variables to allow for up to 8-way analyses within groups for some outcomes at time of final analysis (see supplemental file 1 for more detail).

### Recruitment

Recruitment of children in the target age range began with a first cohort A from May to August 2021. Due to pandemic-related disruptions, recruitment continued with a second cohort B from March to June 2022 for a total of 1495 children and their parents and 1638 teachers enrolled in the study to date. New teachers are being recruited, consented, and trained as they begin employment at BestStart (see supplemental files 1, 3).

### Allocation to conditions (randomization)

The procedure for allocating centres to conditions was central randomization by computer using a 2 (ENRICH vs no ENRICH) x 2 (ENGAGE vs no ENGAGE) factorial design (see Figure 2). The method used to generate the sequence was permuted block randomization by early childhood centre.

### Data collection

In the Best Start main trial, assessments are being administered approximately every 9 months between ages 1.5 and 5 years, and at age 6 years. Teachers and parents are asked to complete instruments to assess children’s oral language and literacy, self-regulation, and socioemotional skills (see Figure 1), which takes approximately 35 minutes each wave using REDCap (Research Electronic Data Capture) hosted at the University of Otago [32, 33]. Paper forms are also provided for participants who do not wish to complete the forms online.

In the Video Project, naturally occurring classroom practices at 24 centres are videotaped by researchers every year to age 5 years, which takes approximately an hour in each centre at each assessment wave.

In the Brain and Behavior Development sub-study, a subset of 237 children from the main trial are taking part in additional neurophysiological and behavioral assessments: EEG/ERP, eye-tracking, and behavioral measures of attention, emotion processing, categorization, cognitive flexibility, inhibitory control, and memory, as well as a parent-child interaction task (see Figure 1; supplemental file 2 for protocol).

Data collection for the Best Start main trial, the Video Project sub-study, and the Brain and Behavior Development sub-study are targeted to end in March 2025.

Incentives to complete assessments at each wave in the main trial are in the form of raffles at each wave for centres (of ten $250 gift cards) and parents (of ten $250 gift cards). Parents and children participating in the Brain and Behavior Development sub-study receive a $20 gift card and small gift at each wave.

### Data management and security

#### Source and format of data

All raw data files including the reports of teachers and parents are saved as CSV files, and media files including the naturalistic observations from 24 BestStart centres are saved as MTS video files at the University of Otago.

#### Data sharing protocol between centres and the university

Data from all sites are collected by the REDcap server or sent directly to the research manager, Dr. Tugce Bakir-Demir, at University of Otago. Hard copies of any paper forms are couriered to the University of Otago in a secure envelope via New Zealand Couriers.

Data sharing agreements have been established between the University of Otago, University of Auckland, and Boston Children’s Hospital (BCH). Participants were informed and consented to the data sharing process (see supplemental file 3).

#### Data storage, security, and access

Raw digital data are stored in REDCap, which is a secure, web-based software platform. Back-ups of the digital data are also on the University of Otago server. Any original non-digital data (including any paper forms and micro-SD cards with video footage) are securely stored at the research laboratory within the university to which only a limited number of approved researchers in the trial have access.

Raw electrophysiological (EEG/eye-tracking) data are stored encrypted on secure servers within the University of Auckland. Due to the nature of this electrophysiological data, it could not be wholly deidentified prior to processing as it contains audio and video recordings of participants, which are naturally identifying.

### Confidentiality

All information that we collect is used only by the research team working on this study. Any raw data (including video data) and personal information will be retained in secure storage for at least 5 years after the end of the project, as required by the University of Otago’s research policy, after which time it will be destroyed. The overall results of the project will be published and will be available in university libraries and public good databases, but each individual participant’s information will remain anonymous and confidential.

### Statistical analyses

For the primary outcomes of the main trial, we will analyze data only from participants who have remained in their original BestStart centre. All enrolled children will be included in analyses of the B4 School Check and IDI. We will use intent-to-treat analyses for the participants in the Brain and Behaviour Development sub-study, who will continue to be followed even if they leave their BestStart centre.

Mixed modelling and moderator/mediator analyses will be conducted to assess children’s oral language and self-regulation outcomes as a function of intervention condition (nested within centres) and as a function of implementation fidelity, adjusting for confounders. Where appropriate, we will use multiple imputation to account for missing data and run sensitivity analyses that compare the results for each cohort (A and B) with the results for the entire sample.

## Ethical Considerations

The proposed study does not involve any medication or invasive procedures and as such risk to children and their families is minimal. The techniques we will teach are designed to be positive and beneficial for adults and children alike; however, conversations can sometimes become negative, and young children can vary in their engagement with new activities. We anticipate that any of these issues will be within the realm of what teachers normally deal with on an everyday basis, and in fact, the techniques are designed to help educators tackle discussions of everyday negative events in ways that will support children’s coping and emotion regulation, and in ways that will facilitate children’s participation in activities. Therefore, we have determined that there is no need for a formal data monitoring committee.

The University of Otago Health Ethics Committee approved this study on November 23, 2020 (H20/116). The cultural responsiveness of project has also been approved twice by the Ngai Tāhu Research Advisory Committee, once in 2020 and again in 2022. The cultural appropriateness of assessments and intervention materials is continually monitored by our own Cultural Advisory Group who are drawn from academia, the community, and the BestStart provider (currently Amanda Clifford, Elizabeth Schaughency, Mele Taumoepeau, Karen Salmon, Pip Laufiso, Barbara Backshall, and Waveney Lord).

## Trial Registration Status

The study was registered in the Australia/New Zealand Clinical Trial Registry (ACTRN12621000845831) on 22 April 2021, prior to the date of the first enrolment on 4 May 2021. Confirmation of trial registration was delayed (1 July 2021) due to a COVID-related backlog.

## Discussion

The Best Start study was developed to promote oral language and self-regulation development in children from 1.5 years of age using interventions that include both teachers and parents. To the best of our knowledge, this study is the first RCT to test the singular and combined effects of oral language and self-regulation interventions on children from such a young age.

Our findings will inform both theory and practice. With respect to theory, we will be able to test our hypothesis that oral language is a key driver of later self-regulation [11]. We will also test our hypothesis that combining oral language and self-regulation interventions will produce the greatest benefits for children’s self-regulation and later academic functioning. Finally, via the Brain and Behavior Development sub-study, we will test our hypothesis that the intervention benefits are mediated by neurophysiological changes in brain development.

With respect to practice, our findings will inform the design of early childhood curricula in New Zealand and internationally. Critically, each intervention is designed so that it can be readily scaled up to reach larger numbers of early childhood teachers and children — within a short-time frame.

## Supporting information

Supplemental File 1 Original Study Protocol

Supplemental File 2 Brain and Behaviour Development Sub-Study Protocol

Supplemental File 3 Information Sheets and Consent Forms

## Data Availability

No datasets were generated or analysed during the current study protocol.

## Dissemination

We will disseminate the findings widely via newsletters to early childhood centres and parents twice a year, in-person and online presentations at national and international conferences, publications in academic journals, blogs and podcasts for professional and trade organizations, oral and written communication to the New Zealand Ministry of Education, and a study website.

## Supplemental Information

- File 1: Original protocol for Main Trial from Ethics application submitted in October 2020
- File 2: Original protocol for Brain and Behavior Development sub-study submitted in February 2021
- File 3: Information sheets and consent forms for the Main Trial and Brain and Behavior Development sub-study

## Acknowledgments

We acknowledge with gratitude all of the BestStart teachers, professional practice leaders, centre managers, and staff who are contributing to the study; and to Julia Errington-Smith, Grace Lam, and other support staff at Methodist Mission Southern. Most of all, we thank the parents and children for their participation.

## Author Contributions

**<u>Conceptualization:</u>** Richie Poulton, Stuart McNaughton, Elaine Reese, Clair Edgeler, Jimmy McLauchlan, Elizabeth Schaughency, Mele Taumoepeau, Karen Salmon, Charles Nelson

**<u>Data curation:</u>** Tugce Bakir-Demir, Hayley Guiney, Natasha Maruariki

**<u>Formal analysis:</u>** Hayley Guiney, Jesse Kokaua, Tugce Bakir-Demir

**<u>Funding acquisition:</u>** Richie Poulton

**<u>Methodology:</u>** Richie Poulton, Elaine Reese, Jesse Kokaua, Charles Nelson, Elizabeth Schaughency, Mele Taumoepeau, Karen Salmon, Amanda Clifford, Ran Wei, Valentina Pergher, Sophia Amjad, Anita Trudgen

**<u>Project administration:</u>** Jimmy McLauchlan, Natasha Maruariki, Richie Poulton, Elaine Reese, Tugce Bakir-Demir, Hayley Guiney, Justin O’Sullivan

**<u>Resources:</u>** Elaine Reese, Elizabeth Schaughency, Mele Taumoepeau, Jimmy McLauchlan, Julia Errington Scott

**<u>Supervision of postgraduate students:</u>** Elaine Reese, Amanda Clifford, Elizabeth Schaughency, Mele Taumoepeau, Tugce Bakir-Demir, Hayley Guiney

**<u>Writing – original draft:</u>** Elaine Reese, Tugce Bakir-Demir, Hayley Guiney

**<u>Writing – review & editing:</u>** Elaine Reese, Richie Poulton, Jesse Kokaua, Hayley Guinea, Tugce Bakir-Demir, Jimmy McLauchlan, Justin O’Sullivan, Clair Edgeler, Elizabeth Schaughency, Mele Taumoepeau, Karen Salmon, Amanda Clifford, Natasha Maruariki, Stuart McNaughton, Peter Gluckman, Charles Nelson, Ran Wei, Valentina Pergher, Sophia Amjad, Anita Trudgen

## References

1. Robson DA, Allen MS, Howard SJ. Self-regulation in childhood as a predictor of future outcomes: A meta-analytic review. Psychological Bulletin. 2020;146(4):324-54. Epub 2020/01/07. doi: 10.1037/bul0000227. PubMed PMID: 31904248.

2. Hendry A, Jones EJH, Charman T. Executive function in the first three years of life: Precursors, predictors and patterns. Developmental Review. 2016;42:1–33. doi: 10.1016/j.dr.2016.06.005.

3. Moffitt TE, Arseneault L, Belsky D, Dickson N, Hancox RJ, Harrington H, et al. A gradient of childhood self-control predicts health, wealth, and public safety. Proceedings of the National Academy of Sciences. 2011;108(7):2693–8. Epub 2011/01/26. doi: 10.1073/pnas.1010076108. PubMed PMID: 21262822; PubMed Central PMCID: PMCPMC3041102.

4. Richmond-Rakerd LS, Caspi A, Ambler A, d’Arbeloff T, de Bruine M, Elliott M, et al. Childhood self-control forecasts the pace of midlife aging and preparedness for old age. PNAS Proceedings of the National Academy of Sciences of the United States of America. 2021;118(3). Epub 2021/01/06. doi: 10.1073/pnas.2010211118. PubMed PMID: 33397808; PubMed Central PMCID: PMCPMC7826388.

5. Blair C. Developmental science and executive function. Current Directions in Psychological Science. 2016;25(1):3–7. Epub 2016/03/18. doi: 10.1177/0963721415622634. PubMed PMID: 26985139; PubMed Central PMCID: PMCPMC4789148.

6. Mohr-Jensen C, Steinhausen HC. A meta-analysis and systematic review of the risks associated with childhood attention-deficit hyperactivity disorder on long-term outcome of arrests, convictions, and incarcerations. Clinical Psychology Review. 2016;48:32–42. Epub 2016/07/09. doi: 10.1016/j.cpr.2016.05.002. PubMed PMID: 27390061.

7. Yang Y, Shields GS, Zhang Y, Wu H, Chen H, Romer AL. Child executive function and future externalizing and internalizing problems: A meta-analysis of prospective longitudinal studies. Clinical Psychology Review. 2022;97:102194. Epub 2022/08/15. doi: 10.1016/j.cpr.2022.102194. PubMed PMID: 35964337.

8. Healey DM, Halperin JM. Enhancing Neurobehavioral Gains with the Aid of Games and Exercise (ENGAGE): Initial open trial of a novel early intervention fostering the development of preschoolers’ self-regulation. Child Neuropsychology. 2015;21(4):465–80. Epub 2014/04/17. doi: 10.1080/09297049.2014.906567. PubMed PMID: 24735230.

9. Healey D, Healey M. Randomized Controlled Trial comparing the effectiveness of structured-play (ENGAGE) and behavior management (TRIPLE P) in reducing problem behaviors in preschoolers. Scientific Reports. 2019;9(1):3497. Epub 2019/03/07. doi: 10.1038/s41598-019-40234-0. PubMed PMID: 30837595; PubMed Central PMCID: PMCPMC6401110.

10. Healey D, Milne B, Healey M. Adaption and implementation of the engage programme: Teaching self-regulation through play, within the early childhood curriculum. Research Square. 2022. doi: 10.21203/rs.3.rs-1359890/v1.

11. Salmon K, O’Kearney R, Reese E, Fortune CA. The role of language skill in child psychopathology: Implications for intervention in the early years. Clinical Child and Family Psychology Review. 2016;19(4):352–67. Epub 2016/10/28. doi: 10.1007/s10567-016-0214-1. PubMed PMID: 27678011.

12. Gilkerson J, Richards JA, Warren SF, Oller DK, Russo R, Vohr B. Language experience in the second year of life and language outcomes in late childhood. Pediatrics. 2018;142(4). Epub 2018/09/12. doi: 10.1542/peds.2017-4276. PubMed PMID: 30201624; PubMed Central PMCID: PMCPMC6192025.

13. Bleses D, Jensen P, Slot P, Justice L. Low-cost teacher-implemented intervention improves toddlers’ language and math skills. Early Childhood Research Quarterly. 2020;53:64–76. doi: 10.1016/j.ecresq.2020.03.001.

14. Dowdall N, Melendez-Torres GJ, Murray L, Gardner F, Hartford L, Cooper PJ. Shared picture book reading interventions for child language development: A systematic review and meta-analysis. Child Development. 2020;91(2):e383–e99. Epub 2019/02/10. doi: 10.1111/cdev.13225. PubMed PMID: 30737957.

15. Salmon K, Reese E. The benefits of reminiscing with young children. Current Directions in Psychological Science. 2016;25(4):233–8. doi: 10.1177/0963721416655100.

16. Clifford A, Schaughency, E., Das, S., Carroll, J., Riordan, J., & Reese, E. Tender Shoots: Effects of a parent conversation program to promote quality reminiscing and children’s socioemotional development. Manuscript in preparation. 2022.

17. Reese E, Barrett-Young A, Gilkison L, Carroll J, Das S, Riordan J, et al. Tender Shoots: A parent book-reading and reminiscing program to enhance children’s oral narrative skills. Reading and Writing. 2022. doi: 10.1007/s11145-022-10282-6.

18. Riordan J, Reese E, Das S, Carroll J, Schaughency E. Tender Shoots: A randomized controlled trial of two shared-reading approaches for enhancing parent-child interactions and children’s oral language and literacy skills. Scientific Studies of Reading. 2021;26(3):183–203. doi: 10.1080/10888438.2021.1926464.

19. Schaughency E, Linney, K., Carroll, J., Das, S., & Riordan, J. & Reese, E. Tender Shoots: Effects of a preschool shared book-reading preventive intervention on children’s reading skills in the first year of school. Manuscript under review. 2022.

20. Duncan GJ, Dowsett CJ, Claessens A, Magnuson K, Huston AC, Klebanov P, et al. School readiness and later achievement. Developmental Psychology. 2007;43(6):1428–46. Epub 2007/11/21. doi: 10.1037/0012-1649.43.6.1428. PubMed PMID: 18020822.

21. Mol SE, Bus AG, de Jong MT, Smeets DJH. Added value of dialogic parent–child book readings: A meta-analysis. Early Education and Development. 2008;19(1):7–26. doi: 10.1080/10409280701838603.

22. Wasik BA, Hindman AH, Snell EK. Book reading and vocabulary development: A systematic review. Early Childhood Research Quarterly. 2016;37:39–57. doi: 10.1016/j.ecresq.2016.04.003.

23. Lonigan CJ, Whitehurst GJ. Relative efficacy of parent and teacher involvement in a shared-reading intervention for preschool children from low-income backgrounds. Early Childhood Research Quarterly. 1998;13(2):263–90. doi: 10.1016/s0885-2006(99)80038-6.

24. Whitehurst GJ, Epstein JN, Angell AL, Payne AC, Crone DA, Fischel JE. Outcomes of an emergent literacy intervention in Head Start. Journal of Educational Psychology. 1994;86:542–55. doi: 10.1037/0022-0663.86.4.542.

25. Reese E, Keegan P, McNaughton S, Kingi TK, Carr PA, Schmidt J, et al. Te Reo Maori: Indigenous language acquisition in the context of New Zealand English. Journal of Child Language. 2018;45(2):340–67. Epub 2017/07/07. doi: 10.1017/S0305000917000241. PubMed PMID: 28679455.

26. Putnam SP, Rothbart MK. Development of short and very short forms of the Children’s Behavior Questionnaire. Journal of Personality Assessment. 2006;87(1):102–12. Epub 2006/07/22. doi: 10.1207/s15327752jpa8701_09. PubMed PMID: 16856791.

27. Dickinson DK, McCabe, A., & Sprague, K. Teacher Rating of Oral Language and Literacy (TROLL): Individualizing early literacy instruction with a standards-based rating tool. The Reading Teacher. 2003;56(6):554–64. doi: https://link.gale.com/apps/doc/A99113524/AONE?u=otago&sid=googleScholar&xid=9fa87c74.

28. Bird A, Reese, E., Schaughency, E., Atatoa Carr, P., & Morton, S. M. B. Talking, praising and teaching: How does parent interaction during a learning task relate to children’s early learning?. Manuscript under review. 2022.

29. Bronson MB, Goodson, B. D., Layzer, J. I., & Love, J. M. Child Behavior Rating Scale: Cambridge, MA: ABT Associates.; 1990.

30. Gibb S, Milne B, Shackleton N, Taylor BJ, Audas R. How universal are universal preschool health checks? An observational study using routine data from New Zealand’s B4 School Check. BMJ Open. 2019;9(4):e025535. Epub 2019/04/06. doi: 10.1136/bmjopen-2018-025535. PubMed PMID: 30948582; PubMed Central PMCID: PMCPMC6500230.

31. Dickinson DK, Collins MF, Nesbitt K, Toub TS, Hassinger-Das B, Hadley EB, et al. Effects of teacher-delivered book reading and play on vocabulary learning and self-regulation among low-income preschool children. Journal of Cognition and Development. 2018;20(2):136–64. doi: 10.1080/15248372.2018.1483373.

32. Harris PA, Taylor R, Minor BL, Elliott V, Fernandez M, O’Neal L, et al. The REDCap consortium: Building an international community of software platform partners. Journal of Biomedical Information. 2019;95:103208. Epub 2019/05/13. doi: 10.1016/j.jbi.2019.103208. PubMed PMID: 31078660; PubMed Central PMCID: PMCPMC7254481.

33. Harris PA, Taylor R, Thielke R, Payne J, Gonzalez N, Conde JG. Research electronic data capture (REDCap)--a metadata-driven methodology and workflow process for providing translational research informatics support. Journal of Biomedical Information. 2009;42(2):377–81. Epub 2008/10/22. doi: 10.1016/j.jbi.2008.08.010. PubMed PMID: 18929686; PubMed Central PMCID: PMCPMC2700030.

