## Supplemental File 1 Original Study Protocol for "The Best Start (Kia Tīmata Pai): A Study Protocol for a Cluster Randomized Trial with Early Childhood Teachers to Support Children’s Oral Language and Self-Regulation Development"

Supplemental File 1
Original Study Protocol Submitted to University of Otago Health Ethics Committee
October 2020

### Background

Self-regulation is a vital skill for people of all ages. Self-regulation in early childhood predicts later academic functioning and life success (Moffitt et al., 2011). Poor self-regulation in early childhood is associated with current behaviour problems and with later psychopathology. For example, ADHD is a chronic neurodevelopmental disorder of self-regulation (Mannuzza et al., 2003; Willcutt et al., 2005), and the most common disorder of childhood (APA, 2000) with 12,500 children medicated for ADHD in New Zealand (New Zealand Health Information Service, 2008). ADHD is associated with increases in the risk of academic and employment failure, additional psychopathology, and criminality (Gathje et al., 2008).

Although *external* interventions using medication (Conners, 2002) and behaviour modification (e.g., parent training; Pelham & Fabiano, 2008) are highly effective treatments for ADHD in the short-term, gains are rarely maintained after the termination of treatment (Molina et al., 2009). Healey and Halperin (2015) developed a novel preschool intervention designed to overcome these shortcomings. ENGAGE (Enhancing Neurocognitive Growth with the Aid of Games and Exercise) aims to improve ADHD symptoms through strengthening neural networks leading to improved self-regulatory skills. ENGAGE employs a range of tasks involving skill areas known to be under-developed in children with ADHD. The tasks are variations of well-known children's games (e.g., Simon Says) that are interpersonal in nature, and teach a range of physical and mental skills. ENGAGE leads to equivalent improvements in parent-rated behaviour problems as a gold-standard parent-management programme (Triple P), with treatment gains maintained 12 months later (Healey & Healey, 2019).

Another way to foster self-regulation is to enhance children's oral language development (Salmon et al., 2016). The way adults talk with children during everyday activities (mealtimes, book-reading, play) advances children's early language and cognitive development (e.g., Gilkerson et al., 2018), which in turn enhances their self-regulation. Oral language skills also support children's literacy development and their success in school. *Tender Shoots* is a book-reading and conversation programme for parents and educators to enhance pre-schoolers' oral language development (Schaughency et al., 2014). *Tender Shoots* stimulates high-quality conversations between adults and children (Das et al., in prep), which in turn improves children's oral language, literacy, self-regulation, and socioemotional skills (Das et al., in prep; Reese et al., 2020; Riordan et al., in prep; Schaughency et al., 2020; in prep).

We now need to know if ENGAGE and *Tender Shoots* together will produce even better outcomes for children than either programme alone. To maximise these benefits, we are trialling a new preparatory phase for toddlers, ENRICH (ENhancing RICH conversations), that will lay the oral language foundations needed for children to make the most of ENGAGE and *Tender Shoots* in the preschool years.

Our aim is to conduct a 5-year longitudinal randomised controlled trial (RCT) evaluating the effectiveness of targeting language and self-regulation over time (ENRICH + *Tender Shoots* + ENGAGE), to targeting language alone (ENRICH + *Tender Shoots*) or self-regulation alone (ENGAGE), in comparison to the usual BestStart curriculum.

The four arms are thus (400 children in each).
1) **Combined**: ENRICH (1.5 to 3 years) + *Tender Shoots* (3 to 5 years) + ENGAGE (3 to 5 years)
2) **Language only**: ENRICH (1.5 to 3 years) + *Tender Shoots* (3 to 5 years)
3) **Self-regulation only**: ENGAGE (3 to 5 years)
4) **Control**: Existing BestStart curriculum
*Note that groups 1, 2, and 3 will also receive the existing BestStart curriculum.

| **Intervention groups** | **Wave 1 1.5 – 3 years** | | **Wave 2 3 – 5 years** | | |
|---|---|---|---|---|---|
| | **ENRICH** | **As usual** | ***Tender Shoots*** | **ENGAGE** | **As usual** |
| **Combined** | **X** | **X** | **X** | **X** | **X** |
| **Language** | **X** | **X** | **X** | | **X** |
| **Self-regulation** | | **X** | | **X** | **X** |
| **Control** | | **X** | | | **X** |

**Hypotheses:** We expect that with its focus on improving self-regulation, ENGAGE will produce the strongest benefits for self-regulation. But we expect the combination of ENRICH, *Tender Shoots,* and ENGAGE will lead to the best outcomes for children's self-regulation and socioemotional skills, because we hypothesise that language development will potentiate the benefits of ENGAGE on children's self-regulation. We will conduct mediator analyses to assess whether benefits are gleaned through increases self-regulation or language. We also expect that the combination of ENRICH + *Tender Shoots*, with its focus on improving language, will indeed produce the greatest benefits for children's language development. These language benefits are expected to extend to children's reading skills and be maintained through the first year of primary school. We expect that either ENGAGE or ENRICH/*Tender Shoots* alone will also lead to benefits for self-regulation and oral language, respectively, over the existing BestStart curriculum. From a policy point of view, it would be of interest to see which intervention gives best value for money, or if the combination of the three is worth the extra cost and time. The findings from this study could shape the national early childhood curriculum.

**Figure 1. Hypothetical Timeline of Study Activities to Start of Wave 2.**

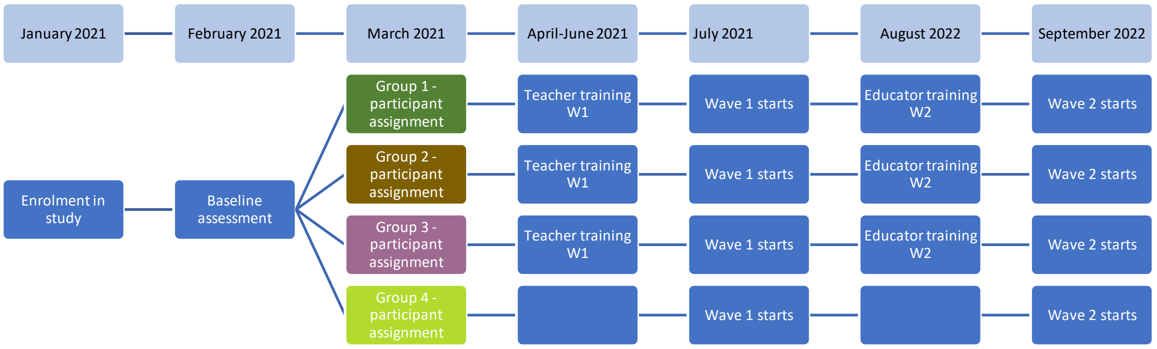


**Sampling, Power, Allocation:** Our methodologist and data analyst, Dr. Matt Healey (Methodist Mission Southern), will select 140 centres from over 200 BestStart centres to form a nationally representative sample of regions, socioeconomic status, and children's ethnicity (see attached methodology document). This number of centres is estimated to allow recruitment of the desired number of 1600 children aged 17- to 24-months at the outset. A final sample size of 400 children per group is estimated to provide sufficient power to mitigate the possible effects of cluster variance confounds, interaction effects, predictor variables, and participant attrition. Dr. Healey will then randomly assign each centre to one of the four conditions.

**Recruitment:** Teachers in each centre will recruit parents to enrol their 17- to 24-month-old children in the study. Children with a pervasive developmental disorder or history of brain trauma will be excluded from data collection. We will thus be unable to generalise to children with pervasive developmental disorders or history of brain trauma. All children within the target age group will be exposed to the interventions within their ECE, but data will not be collected on those children whose families have not granted consent. Parents of nonparticipating children attending the centre will be sent an information letter to let them know about the projects and about their children's educators' participation. There will be no gender, racial/ethnic, language, or socioeconomic restrictions to participation in this study.

**Baseline**: Following enrolment in the study and informed consent, parents and teachers will be asked to complete some of the following measures, which will be decided upon in collaboration with BestStart to minimise response burden: the Behavioural Assessment System for Children (BASC-2; a widely used measure of psychosocial functioning; see Appendix 1); the Strengths and Difficulties Questionnaire (SDQ; see Appendix 2), the Children's Behaviour Questionnaire

(CBQ; see Appendix 3), and the MacArthur New Zealand Communicative Development Inventories in all of the children's languages (NZCDI; Appendix 4). Parents will also be asked to complete a demographic and developmental history questionnaire (see Appendix 5) and the Parenting Stress Index (see Appendix 6) as a baseline measure of stress in relation to parenting their target child, and to report on their tobacco/alcohol/drug use during pregnancy. Teachers will report on the children's skills using the Teacher Rating of Oral Language and Literacy (TROL; see Appendix 7) and the Child Behaviour Rating Scale (CBRS; see Appendix 8). Teachers will also report on their beliefs and current practices to support children's self-regulation, language, and (Brackett et al., 2012). We will offer parents and teachers $10 vouchers for each wave of questionnaires that they are asked to complete. We will give children small rewards (stamps, stickers) at individual assessment sessions.

**Intervention Phase:**
After clusters of centres are randomly assigned to one of the four conditions (with approximately 400 children each), the PIs will hold professional development sessions with 14 professional practice leaders (PPLs) serving around 20 centres each (of which roughly 10 centres for each PPL will be part of the RCT). The PIs will train the PPLs to train the teachers in their participating centres via a training video and resources. A separate training video will be developed for any parents who want to learn the techniques to use at home; this variable will be controlled for in analyses by measuring the number of parents who request the training video and who report using the techniques at home. For the control condition, measures of parents' existing use of the techniques will be taken. These training sessions will be held in early 2021 for ENRICH, and in 2022 for ENGAGE and *Tender Shoots*. We will measure implementation through observations of classroom activities, which are expected to be conducted on a daily basis. We will guard against cross-contamination by taking advantage of the existing structure of the BestStart organisation, in which each PPL manages a cluster of centres. Each PPL will be responsible for administering a single condition across their centres, thus limiting the possibility of teachers or parents sharing information about other conditions with other centres. We will also liase with PPLs throughout the study on ways to prevent cross-contamination across centres in the same region.

**Early Childhood Outcomes**:
Our main outcome measures will be language, self-regulation, socioemotional, and early literacy development. Approximately every 6 months between ages 2 and 5, we will ask teachers and parents to use the same instruments to assess children's self-regulation, language, and socioemotional skills. We will supplement these parent- and teacher-report instruments with individually administered assessments to children from age 2.5, such as the following well-validated measures that co-PIs Healey, Reese, and Schaughency have used in previous projects (e.g., project 16/016):

**Oral language and literacy:** Clinical Evaluation of Language Fundamentals, Australian/New Zealand 5th edition: Preschool 2 (CELF:P; Wiig et al., 2017), the Preschool Early Literacy Indicators (PELI, Kaminski et al., 2014), and children's narrative comprehension and production (Reese et al., 2010). For bilingual children, we will conduct measures of early literacy and narrative in both languages as often as possible.

**Self-regulation:** The Statues subtest from the Developmental Neuropsychological Assessment (NEPSY-2; Korkman et al., 2007); the Head and Toes Task (Ponitz et al., 2008)**;** selected subtests from the Stanford Binet (SB5; Roid, 2003; e.g., Working Memory); and additional subtests from the Developmental Neuropsychological Assessment (NEPSY-2) such as Comprehension of Instructions and Visuomotor Precision.

**Socioemotional skills:** Emotion knowledge task (Denham, 2006); Challenging Behaviour Task (Bierman et al., 2014).

**B4 School Check**: Not all parents obtain a B4 School Check for their children with GPs (Schluter et al., 2020), but we will request access to this information when available. The B4 School Check is a health and development screening with 4-year-olds that assesses communication and behavioural difficulties. This information will serve as a reference point when comparing the children in our sample to other New Zealand children.

**School Outcomes**: We will invite children's primary school teachers to complete the same behavioural, socioemotional, and language and literacy rating scales as the early childhood teachers. If we have the capacity, we will also administer individual assessments, similar to the PELI above, to assess children's developing literacy skills.

### Internal Data Safety Monitoring Committee

We will coordinate with BestStart to develop a protocol for monitoring any adverse events. This protocol will involve Methodist Mission Southern regularly checking in on a weekly basis with individual educators and parents to answer questions and provide support, in collaboration with BestStart management and with the study team.

### References

American Psychiatric Association (2000). *Diagnostic and Statistical Manual of Mental Disorders, Fourth Edition*. Washington, D.C.: American Psychiatric Association.

Bierman, K. L., Nix, R. L., Heinrichs, B. S., Domitrovich, C. E., Gest, S. D., Welsh, J. A., & Gill, S. (2014). Effects of Head Start REDI on children's outcomes 1 year later in different kindergarten contexts. *Child Development*, *85*(1), 140–159. https://doi.org/10.1111/cdev.12117

Brackett, M. A., Reyes, M. R., Rivers, S. E., Elbertson, N. A., & Salovey, P. (2012). Assessing teachers' beliefs about social and emotional learning. *Journal of Psychoeducational Assessment*, *30*(3), 219–236.

https://doi.org/10.1177/0734282911424879

Conners, C, K. (2002). Forty years of methylphenidate treatment in Attention-Deficit /Hyperactivity disorder. *Journal of Attention Disorders, 6*, 17–30.

Das, S., Schaughency, E., Riordan, J., Carroll, J., & Reese, E. (in prep). *Tender Shoots*: Fostering parent-child conversations nurtures children's socioemotional skills.

Denham, S. (2006). The answer is readiness: Now what is the question ? *Early Education and Development*, *17*(1), 57–89. https://doi.org/10.1207/s15566935eed1701

Gathje, R. A., Lewandowski, L. J., & Gordon, M. (2008). The role of impairment in the diagnosis of ADHD. *Journal of Attention Disorders*, *11*(5), 529–537. https://doi.org/10.1177/1087054707314028

Gilkerson, J., Richards, J. A., Warren, S. F., Oller, D. K., Russo, R., & Vohr, B. (2018). Language experience in the second year of life and language outcomes in late childhood. *Pediatrics*, *142*(4). https://doi.org/10.1542/peds.2017-4276

Healey, D. M., & Halperin, J. M. (2015). Enhancing neurobehavioral gains with the aid of games and exercise (ENGAGE): Initial open trial of a novel early intervention fostering the development of preschoolers self-regulation. *Child Neuropsychology*, *21*(4), 465–480. https://doi.org/10.1080/09297049.2014.906567

Healey, D., & Healey, M. (2019). Randomized Controlled Trial comparing the effectiveness of structured-play (ENGAGE) and behavior management (TRIPLE P) in reducing problem behaviors in preschoolers. *Scientific Reports*, *9*(1), 1–9. https://doi.org/10.1038/s41598-019-40234-0

Kaminski, R. A., Abbott, M., Bravo Aguayo, K., Latimer, R., & Good, R. H. (2014). The Preschool Early Literacy Indicators: Validity and benchmark goals. *Topics in Early Childhood Special Education*, *34*(2), 71–82. https://doi.org/10.1177/0271121414527003

Korkman, M., Kirk, U., & Kemp, S. (2007). *NEPSY-II: Clinical and interpretive manual*. The Psychological Corporation.

Mannuzza, S., Klein, R. G., & Moulton, J. L. (2003). Persistence of attention-deficit/hyperactivity disorder into adulthood: What have we learned from the prospective follow-up studies? *Journal of Attention Disorders*, *7*(2), 93–100. https://doi.org/10.1177/108705470300700203

Moffitt, T. E., Arseneault, L., Belsky, D., Dickson, N., Hancox, R. J., Harrington, H. L., Houts, R., Poulton, R., Roberts, B. W., Ross, S., Sears, M. R., Thomson, W. M., & Caspi, A. (2011). A gradient of childhood self-control predicts health, wealth, and public safety. *Proceedings of the National Academy of Sciences of the United States of America*, *108*(7), 2693–2698. https://doi.org/10.1073/pnas.1010076108

Molina, B. S. G., Hinshaw, S. P., Swanson, J. M., Arnold, L. E., Vitiello, B., Jensen, P. S., Epstein, J. N., Hoza, B., Hechtman, L., Abikoff, H. B., Elliott, G. R., Greenhill, L. L., Newcorn, J. H., Wells, K. C., Wigal, T., Gibbons, R. D., Hur, K., & Houck, P. R. (2009). The MTA at 8 years: Prospective follow-up of children treated for combined-type ADHD in a multisite study. *Journal of the American Academy of Child and Adolescent Psychiatry*, *48*(5), 484–500. https://doi.org/10.1097/CHI.0b013e31819c23d0

Pelham, W. E., & Fabiano, G. A. (2008). Evidence-based psychosocial treatments for attention-deficit/hyperactivity disorder. In *Journal of Clinical Child and Adolescent Psychology* (Vol. 37, Issue 1). https://doi.org/10.1080/15374410701818681

Pharmaceutical Information Database (2008). New Zealand Health Information Service. http://www.nzhis.govt.nz/moh.nsf/pagesns/485?Open

Ponitz, C. C. E., McClelland, M. M., Jewkes, A. M., Connor, C. M. D., Farris, C. L., & Morrison, F. J. (2008). Touch your toes! Developing a direct measure of behavioral regulation in early childhood. *Early Childhood Research Quarterly*, *23*(2), 141–158. https://doi.org/10.1016/j.ecresq.2007.01.004

Reese, E., Leyva, D., Sparks, A., Grolnick, W. (2010). Maternal elaborative reminiscing increases low-income children's narrative skills relative to dialogic reading. *Early Education and Development, 21*(3), 318-342. https://doi-org.ezproxy.otago.ac.nz/10.1080/10409289.2010.481552

Reese, E., Macfarlane, L., McAnally, H., Robertson, S. J., & Taumoepeau, M. (2020). Coaching in maternal reminiscing with preschoolers leads to elaborative and coherent personal narratives in early adolescence. *Journal of Experimental Child Psychology*. https://doi.org/10.1016/j.jecp.2019.104707

Robson, D. A., Allen, M. S., & Howard, S. J. (2020). Self-regulation in childhood as a predictor of future outcomes: A meta-analytic review. *Psychological Bulletin*, *146*(4), 324-354. https://doi.org/10.1037/bul0000227

Riordan, J., Reese, E., Das, S., Carroll, J., & Schaughency, E. (in prep). *Tender Shoots:* Helping parents to have rich conversations with young children to advance early literacy.

Roid, G. H. (2003). *Stanford-Binet Intelligence Scales, Fifth Edition: Technical manual*. Riverside Publishing.

Salmon, K., O'Kearney, R., Reese, E., & Fortune, C. A. (2016). The role of language skill in child psychopathology: Implications for intervention in the early years. *Clinical Child and Family Psychology Review*, *19*(4), 352–367. https://doi.org/10.1007/s10567-016-0214-1

Schaughency, E., Das, S., Carroll, J., Johnston, J., Robertson, S.-J., & Reese, E. (2014). Getting ready for school: Exploring caregivers' implementation of a parent-mediated school readiness programme. *7th Educational Psychology Forum*.

Schaughency, E., Linney, K., Carroll, J., Das, S., & Riordan, J. & Reese, E. (in prep). *Tender Shoots*: Effects of a preschool shared book-reading preventive intervention on children's reading skills in the first year of school.

Schaughency, E., Riordan, J., Reese, E., Derby, M., & Gillon, G. (2020). Developing a Community-Based Oral Language Preventive Intervention. *Infants & Young Children*, *33*(3), 195–218. https://doi.org/10.1097/IYC.0000000000000171

Schluter, P. J., Audas, R., Kokaua, J., McNeill, B., Taylor, B., Milne, B., & Gillon, G. (2020). The efficacy of preschool developmental indicators as a screen for early primary school-based literacy interventions. *Child Development, 91*(1), e59-e76.

Wiig, E. H., Semel, E., & Secord, W. A. (2017). *Clinical Evaluation of Language Fundamentals Australian and New Zealand Fifth Edition*. Pearson.

Willcutt, E. G., Doyle, A. E., Nigg, J. T., Faraone, S. V., & Pennington, B. F. (2005). Validity of the executive function theory of attention-deficit/ hyperactivity disorder: A meta-analytic review. *Biological Psychiatry*, *57*(11), 1336–1346. https://doi.org/10.1016/j.biopsych.2005.02.006
