## Supplemental File 2 Brain and Behaviour Development Sub-Study Protocol for "The Best Start (Kia Tīmata Pai): A Study Protocol for a Cluster Randomized Trial with Early Childhood Teachers to Support Children’s Oral Language and Self-Regulation Development"

**Supplemental File 3**

**Original Protocol for Brain Development Sub-Study, Toddler Phase**

1. EEG/ERP Tasks (3 tasks; ~ 30 minutes)
2. Eyetracking (2 tasks; ~ 15 minutes)
3. Behavioral executive function battery (3 tasks; ~15 minutes)
4. Parent- child interaction task (15 minutes)

**Task Descriptions:**

1. **EEG/ERP** (~ 30 minutes total, allowing 5-10 minutes for netting)
   a. **Resting state EEG** (~5 minutes)
      i. EEG is recorded, for 5 minutes, while children watch a video of a screensaver or moving toys (as a distraction)
   b. **Flanker ERP task** (~8 minutes)
      i. Passive ERP task testing attention, discrimination, novelty detection, and information processing. Stimuli are 5 fish in a row. In "congruent" trials, all of the fish are facing in the same direction". In "incongruent" trials, one of the five fish is facing in the opposite direction. Eye gaze data will be collected with eye trackers to see how long it takes for children to detect (look at) the fish facing the opposite direction
   c. **Auditory familiar/non-familiar ERP task** (~8 minutes)
      i. Passive ERP task testing attention, discrimination, detection of novelty, and information processing. Stimuli are familiar voices and non-familiar voices. (details still being thought through - familiar voices could be a recording of the mom's voice. If not possible, a training phase in which the child is familiarized to a voice could be used.

2. **Eyetracking** (~15 minutes, allowing a few minutes for set-up and calibration)
   a. **Disengagement** (~5 minutes)
      i. In each trial, children are presented with a face as the central stimulus. A peripheral stimulus (a geometric shape) is presented to the left or right of the central stimulus (face) with a ~1000 ms delay of onset. There will be a familiar face stimuli (same face for ~70% of trials), and unfamiliar faces (3 different faces - presented 10% of the trials each). The length of time that the children take to shift their eye gaze from the central face target to look at the peripheral target will be measured with the eye tracker.
      ii. **Working memory: (**~5 minutes**)** 4 boxes task (e.g., Garon et al., 2014) with eye tracking. 1 of 4 doors on screen has a toy behind it. Flap goes down. Flap comes back up, does the child look at the correct door?

3. **Behavioral executive functioning battery** (~15 minutes)
    a. **Glitter wand task** (Devine, Ribner & Hughes, 2019) - attractive toy placed within reach of child; Experimenter tells child not to touch it and then turns around. Time how long the child can resist touching (up to 30 seconds). Testing behavioral inhibition.
    b. **Reverse categorisation** - two sets of objects, each with a different color. There are two boxes, one with each of the colors. Children are given objects (e.g. blocks) one at a time and instructed to place them in the box with the *opposite color* as the block. Testing cognitive flexibility.
    c. **Spin the pots task** - cups are arranged on a spinning round disk (lazy susan). Experimenter places stickers under all cups except for 1 or 2 while child is watching; covers cups and spins, then uncovers. Child sequentially picks cups that they think have a sticker under them. Testing working memory.

4. **Parent-child interaction task** (~ 15 minutes)

- No toy play (4 mins)
- Books + toys free play (4 mins)
- Divided attention (parent watches an instructional video while the child plays by him/herself) (developed by Ran Wei; 4 mins)
- Parent-child narrative task (Reese & Newcombe, 2007; talking about a pre-selected shared past event) (3-4 mins)
