## Supplemental File 3 Information Sheets and Consent Forms for "The Best Start (Kia Tīmata Pai): A Study Protocol for a Cluster Randomized Trial with Early Childhood Teachers to Support Children’s Oral Language and Self-Regulation Development"

**INFORMATION SHEET FOR PARTICIPANTS (PARENTS/WHANAU)**

Thank you for your interest and consideration of possible participation for you and your child in our project. Please read this information sheet carefully before deciding whether to participate. If you decide to participate, we thank you. If you decide not to take part, there will be no disadvantage to you, and we thank you for considering our request.

#### What is the aim of the project?

The aim of this project is to discover the "best start" for young children in early childhood education. This research is a 4-year longitudinal study to compare three new evidence-based professional learning and development (PLD) modules for early childhood kaiako/educators, added to the existing BestStart curriculum. The PLD modules integrate effective techniques for developing children's oral language, literacy, self-regulation, and social-emotional competence, all of which are vital for academic achievement. The programme is expected to contribute to kaiako/educators' professional learning and development and to be beneficial for children's development. Your child's centre may be allocated to receive one of the new PLD modules, or to continue to teach, if in one of the important control groups, in the existing BestStart curriculum.

This project is funded by the Wright Family Foundation. It is a collaborative project between BestStart, Methodist Mission Southern and researchers at the University of Otago, Victoria University Wellington, and University of Auckland.

#### What Type of Participants are Being Sought?

Participants will be educators and children in their care aged between 17 months and 5 years, and their parents. Your child's kaiako/educators are also being invited to participate.

#### What will participants be asked to do?

Should you agree to take part in this project with your child, you will be asked to complete questionnaires about your child's health and development, and about your own health, every 6 months until your child turns 5 years old. These questionnaires are expected to take a total of 3 hours across the next 3.5 years. If your child's centre is selected to learn the new techniques, your child's centre will be sharing these practices so that you can learn how to use them at home. To acknowledge your contribution to the project, a small token of appreciation will be offered. Your child may be videotaped as part of our evaluation of classroom practices.

Please be aware that you may decide not to take part in the project without any disadvantage to yourself or your child.

#### What data or information will be collected and what use will be made of it?

In addition to the information described above, we will also collect general demographics about you and your family (age, ethnicity, gender etc.). The purpose of collecting demographic information is so that we may describe our study sample and further tailor the programme to the needs of individual families. We will also seek access to your child's B4 School Check data (Ministry of Health) and the Integrated Data Infrastructure (IDI; Statistics New Zealand) to add to the information we are able to collect in the centre

All information that we collect will be used only by the research team working on this study. The overall results of the project may be published and will be available in the University library, but each individual participant's information will remain anonymous and confidential as described below. You are most welcome to request a copy of the results of the project should you wish.

The data collected will be securely stored in such a way that only the research team will be able to gain access to it. Any raw data and personal information (including video data) will be retained in secure storage for at least five years after the end of the project, as required by the University's research policy, after which time it will be destroyed.

#### Can participants change their mind and withdraw from the project?

Reminder: You and your child may withdraw from participation in the project at any time and without any disadvantage to yourself of any kind

#### What if participants have any questions?

If you have any questions about our project, either now or in the future, please feel free to contact any of the following:


Professor Elaine Reese
(03) 479-8441

Professor Richie Poulton
(03)479-8507

A/P Dione Healey
(03)479-7620

Dr Elizabeth Schaughency
(03)479-5864


This study has been approved by the University of Otago Human Ethics Committee. If you have any concerns about the ethical conduct of the research, you may contact the Committee through the Human Ethics Committee Administrator (ph(03) 479 8256 or). Any issues you raise will be treated in confidence and investigated and you will be informed of the outcome.

**Kia Timata Pai: Fostering Children's Oral Language and Self-Regulation**

CONSENT FORM FOR PARTICIPANTS (PARENTS/WHAN AU)

I have read the Information Sheet concerning this project and understand what it is about. All my questions have been answered to my satisfaction. I understand that I am free to request further information at any stage.

I know that:

1. My participation (and my child's) in the project is entirely voluntary

2. My child and I are free to withdraw from the project at any time without any disadvantage

3. My child will be assessed every 6 months to age 5 for oral language, self-regulation, and social-emotional skills

4. My child may be videotaped as part of the evaluation of classroom practices

5. I consent to the research team contacting my child's primary school after enrolment for a follow-up assessment at age 6

6. I consent to the research team requesting access to my child's B4 School Check data (Ministry of Health)

9. Information collected in this study may be used for future research. This would include comparing my information and my child's with future information in the Integrated Data Infrastructure (IOI). No individualised results will be used in any reports.

10. The results of the project may be published in media and available in the University of Otago Library (Dunedin, New Zealand) but information will be stored and presented in ways that will protect participants' confidentiality.

I agree to take part in this project.

(Signature of participant) (Date)

(Child's name) (Date of birth)

Phone number: ..........................................................................

Address

Street name:

Street number:

Suburb:

City:

Postcode:

This study has been approved by the University of Otago Human Ethics Committee. If you have any concerns about the ethical conduct of the research, you may contact the Committee through the Human Ethics Committee Administrator (ph (03) 479 8256 or). Any issues you raise will be treated in confidence and investigated and you will be informed of the outcome.

### Best Leap

##### A Sub-study to The Best Start Study: Children's Brain Development

##### INFORMATION SHEET FOR PARTICIPANTS (PARENTS)

Thank you for considering participating in this sub-study to our larger project to measure children's brain development. Please read this information sheet carefully before deciding whether or not to participate. If you decide to participate we thank you. If you decide not to take part, there will be no disadvantage to you, and we thank you for considering our request.

**What is the Aim of the Project?**

The aim of this sub-study is to measure the effects of the new practices on children's brain development and their behaviour. We would like to learn more about how the program changes children's attention, cognitive, and social processing.

This project is funded by the international organisation Wellcome LEAP, with collaborators at the Liggins Institute at the University of Auckland, and Boston Children's Hospital in the U.S.

**What Type of Participants are Being Sought?**

Participants will be children who are already participating in the larger BestStart study between 17 months and 36 months, and their parents.

**What will Participants be Asked to Do?**

Should you agree to take part in this project with your child, you will be given taxi or petrol vouchers to visit the Liggins Institute at University of Auckland, or a location closer to your home, for one session every 6 months at times that are convenient for you and your child. Each session will take about one hour. At each session, your child will complete a brain-based measure to assess their neural responses to images and a series of behavioural tasks.

Your child will first look at images on a screen and we will record their eye movements using an eye tracker and their brain activity using electroencephalography (EEG) to record event-related potentials (ERPs). ERPs allow us to measure electrical brain responses to different images.

The eye tracker is made up of a special computer monitor that has a set of infrared cameras built into the edges of the screen. These cameras follow eye movements and will tell us exactly where on the screen your child is looking as they watch the images. In addition, we will record your child's brain activity with a small cap that is made of stretchy material. Each cap has many sponges on it and inside each sponge is a small recording sensor. We soak the cap in a warm salt water solution so the sponges get soft before we put the cap on the child's head. As your child's brain is working, it is constantly giving off small electrical signals, which travel out to the scalp where we can pick them up with the special sensors.

After the eye-tracking and EEG equipment is ready to record, we will have your child sit in front of a computer screen. We will have your child watch a video of moving objects for two minutes while we record their resting brain activity. Next, we will record your child's eye movements and brain activity in response to a series of images on the screen (for instance, rows of fish with one fish facing the wrong way, or a series of faces). In addition, a digital video will be recorded to help the researcher know when to present new images to your child and to aid data analysis. Your child's name will not be associated with the video recording and the file will be accessible only to the investigators of this study. Before starting the behavioural tasks, we will remove the EEG cap from your child's head and take a short break if necessary.

The last three tasks are behavioural measures of your child's executive functioning. To measure waiting for a turn, a researcher will place a glitter wand within reach of your child. The researcher will tell your child not to touch it and then will turn around. We will time how long your child can resist touching (up to 30 seconds). To test cognitive flexibility, your child will be shown two sets of blocks, each with a different colour. They are then shown two boxes. Children are given blocks one at a time and instructed to place them in the box with the *opposite colour* as the block. To test working memory, cups are arranged on a spinning round disk. A researcher places stickers under all cups except for 1 or 2 while your child is watching. Then the researcher covers the cups and spins, then uncovers. Your child will pick cups that they think have a sticker under them.

Please be aware that you may decide not to take part in the project without any disadvantage to yourself of any kind. Even if you decide not to participate in this sub-study, your child can still be part of the larger study.

**What Data or Information will be Collected and What Use will be Made of it?**

All information that we collect will be used only by the research team working on this study. This includes the University of Otago, the Liggins Institute at the University of Auckland, and Boston Children's Hospital in the U.S. We will transfer the data (including video recordings) to Boston Children's Hospital for analysis using secure systems. The overall results of the project may be published and will be available in the University library, but individual participants' information will remain anonymous and confidential as described below. You are most welcome to request a copy of the results of the project should you wish.

The data collected will be securely stored in such a way that only the research team will be able to gain access to it. At the end of the project any personal information (including video recordings) will be destroyed immediately, except that, as required by the University's research policy, any raw data on which the results of the project depend will be retained in secure storage at the University of Otago for at least five years.

**What are the risks of this research study? What could go wrong?**

There are minimal risks posed by these procedures. The study is for research purposes only.

The EEG/ERP technique is a non-invasive technique. If at any time during the session the researcher sees something that you and your doctor should know about, you will be notified and encouraged to see your child's GP. A researcher will be present with you and your child throughout the testing session and the study will be stopped if your child shows any signs of discomfort. The salt solution that we use is non-toxic and will not hurt your child. We will remove most or all of the salt water with a warm flannel before you leave the lab, but some residual salt may remain until your child has a bath. The sensors we use to record brain activity are held together by a stretchy elastic material which may spring back if your child pulls on it. To ensure that this does not happen, a researcher will sit next to your child during the session and will make sure that their hands are not near the cap.

**Can Participants Change their Mind and Withdraw from the Project?**

Reminder: You and your child may withdraw from participation in the project at any time and without any disadvantage to yourself of any kind.

**What if Participants have any Questions?**

If you have any questions about our project, either now or in the future, please feel free to contact any of the following:

**Best Leap Sub-study:**

Sophia Amjad – Research Coordinator
Sonia Byrne – Research Assistant
Anita Trudgen – Research Assistant
The Liggins Institute

**The Best Start Study:**

Professor Elaine Reese
Department of Psychology
University Telephone
479-8441

Professor Richie Poulton
Department of Psychology
University Telephone
479-8507

Associate Professor Dione Healey
Department of Psychology
University Telephone
479-7620

Dr Elizabeth Schaughency
Department of Psychology
University Telephone
479-5864

This study has been approved by the University of Otago Human Ethics Committee. If you have any concerns about the ethical conduct of the research you may contact the Committee through the Human Ethics Committee Administrator (ph +643 479 8256 or). Any issues you raise will be treated in confidence and investigated and you will be informed of the outcome.

### Best Leap

##### A Sub-study to The Best Start: Children's Brain Development

CONSENT FORM FOR
PARTICIPANTS (PARENTS/GUARDIANS)

I have read the Information Sheet concerning this project and understand what it is about. All my questions have been answered to my satisfaction. I understand that I am free to request further information at any stage.

I know that:-

1. My participation (and my child's) in the project is entirely voluntary;

2. My child and I are free to withdraw from the project at any time without any disadvantage;

3. My child's brain development will be assessed every 6 months from 1.5 years (18 months) to 3 years (36 months).

4. I agree for my child to be fitted with the EEG cap.

5. I agree for my child's eye movements to be videotaped.

6. The data (including video recordings) will be transferred securely to Boston Children's Hospital for analysis.

7. Personal identifying information (including video recordings) will be destroyed at the conclusion of the project but any raw data on which the results of the project depend will be retained in secure storage at the University of Otago for at least five years.

8. A small token of appreciation will be offered for my participation.

9. The results of the project may be published and available in the University of Otago Library (Dunedin, New Zealand) but information will be stored and presented in ways that will protect participant's confidentiality.

I agree to take part in this project.

............................................................................ ...............................

(Signature of parent/ guardian) (Date)

............................................................................ ...............................

(Child’s name) (Date of birth)

Phone number: ............................................................................

Address

Street name: ............................................................................

Street number: ............................................................................

Suburb: ............................................................................

City: ............................................................................

Postcode: ............................................................................

This study has been approved by the University of Otago Human Ethics Committee. If you have any concerns about the ethical conduct of the research you may contact the Committee through the Human Ethics Committee Administrator (ph +643 479 8256 or). Any issues you raise will be treated in confidence and investigated and you will be informed of the outcome.
